# High Inspired CO_2_ Target Accuracy in Mechanical Ventilation and Spontaneous Breathing Using the Additional CO_2_ Method

**DOI:** 10.1101/2023.11.19.23298740

**Authors:** Gustav Magnusson, Charalampos Georgiopoulos, Gunnar Cedersund, Lovisa Tobieson, Maria Engström, Anders Tisell

## Abstract

Cerebrovascular Reactivity Imaging (CVR) is a diagnostic method for assessment of alterations in cerebral blood flow in response to a controlled vascular stimulus. The principal utility is the capacity to evaluate the cerebrovascular reserve, thereby elucidating autoregulatory functioning. Over the past decade, CVR has accumulated large interest, emerging as an expanding research field and application in a diverse spectrum of patient populations. In CVR, CO_2_ gas challenge is the most prevalent method, which elicits a vascular response by alterations in inspired CO_2_ concentrations. While several systems have been proposed in the literature, only a limited number have been devised to operate in tandem with mechanical ventilation, thus constraining the majority CVR investigations to spontaneous breathing individuals. We have developed a new method, denoted Additional CO_2_, designed to enable CO_2_ challenge in ventilators. The central idea is the introduction of an additional flow of highly concentrated CO_2_ into the respiratory circuit, as opposed to administration of the entire gas mixture from a reservoir. By monitoring the main respiratory gas flow emanating from the ventilator, the CO_2_ concentration in the inspired gas can be manipulated by adjusting the proportion of additional CO_2_. We evaluated the efficacy of this approach in controlled settings: 1) in a ventilator coupled with a test-lung and 2) in spontaneous breathing healthy volunteers. Additionally, we made a comparative analysis using a conventional method employing a gas reservoir containing a blend of O_2_, N_2_, and CO_2_ in varying concentrations. The methods were evaluated by assessment of the precision in attaining target inspired CO_2_ levels and examination of their performance within a Magnetic Resonance Imaging (MRI) environment. Our investigations revealed that the Additional CO_2_ method consistently achieved a high degree of accuracy in reaching target inspired CO_2_ levels in both mechanical ventilation and spontaneous breathing. We anticipate that these findings will lay the groundwork for a broader implementation of CVR assessments in mechanically ventilated patients.

## 1 INTRODUCTION

Cerebrovascular Reactivity Imaging (CVR) represents an innovative approach for the non-invasive exploration of cerebral hemodynamics. It involves the application of a vasoactive stimulus and simultaneous measurements of alterations in cerebral blood flow. The reactivity, quantified as the change in blood flow divided by the applied stimulus, serves as an indirect indicator of the local vasoregulatory reserve within the cerebral vasculature. Furthermore, this method enables the computation of the time delay in the blood flow response. Research has extensively examined the application of the CVR technique across various medical conditions, including arterial stenosis, moyamoya disease, brain tumors, dementia, small vessel disease, and subarachnoid hemorrhage (Sleight et al., 2021). Despite the promising clinical potential of CVR in these diverse patient cohorts, it has not yet achieved widespread clinical adoption and remains predominantly a research tool. One key impediment to its broader utilization is the limited availability of commercial products for stimulus generation that can be applied across different clinical scenarios.

The established vascular stimulus in CVR measurement is a controlled alteration of arterial carbon dioxide content (aCO_2_). The associated changes in blood flow are typically monitored using magnetic resonance imaging (MRI) in conjunction with the blood oxygen level dependent (BOLD) signal (Liu et al., 2019). Various methods can be employed to manipulate aCO_2_ content, such as controlled breathing patterns, including deep breathing and breath-holding, or the administration of vasoactive drugs like Acetazolamide. However, the preferred approach, due to its reliability and reproducibility, is the administration of carbon dioxide within the inspired gas (Fierstra et al., 2013). Several systems described in the literature use reservoirs with a variable mixture of CO_2_, O_2_, and N_2_, to target different CO_2_ concentrations in the inspired gas (Tancredi et al., 2014; Lu et al., 2014). More sophisticated systems incorporate advanced controls, such as dynamic end-tidal forcing or prospective end-tidal targeting, which enable precise targeting of subjects’ end-tidal CO_2_/O_2_ levels, reflecting the gas concentrations in the alveoli (Wise et al., 2007; Slessarev et al., 2007).

While the literature contains substantial information on systems for CO_2_ gas challenge in spontaneous breathing patients, there has been limited exploration in mechanically ventilated patients (Winter et al., 2010; Venkatraghavan et al., 2018). This gap in research may explain why CVR studies in mechanically ventilated patients have primarily focused on breathing pattern alterations (Brauer et al., 1998; Fierstra et al., 2017; Sari et al., 1990). The aim of this study was to implement a system capable of administering CO_2_ to both ventilated and non-ventilated patients. In contrast to other CO_2_ administration methods, our system does not generate the entire gas mixture but, instead, supplements the inspired gas with additional CO_2_ in proportion to the respiratory gas flow, as illustrated in figure 1, drawing inspiration from nitric oxide systems (Branson et al., 2018). We conducted tests of our system within a ventilator setup alongside a test-lung and in healthy volunteers, comparing it to a conventional CO_2_ gas challenge system, inspired by the approach detailed by Tancredi et al. (2014).

**Figure 1.**
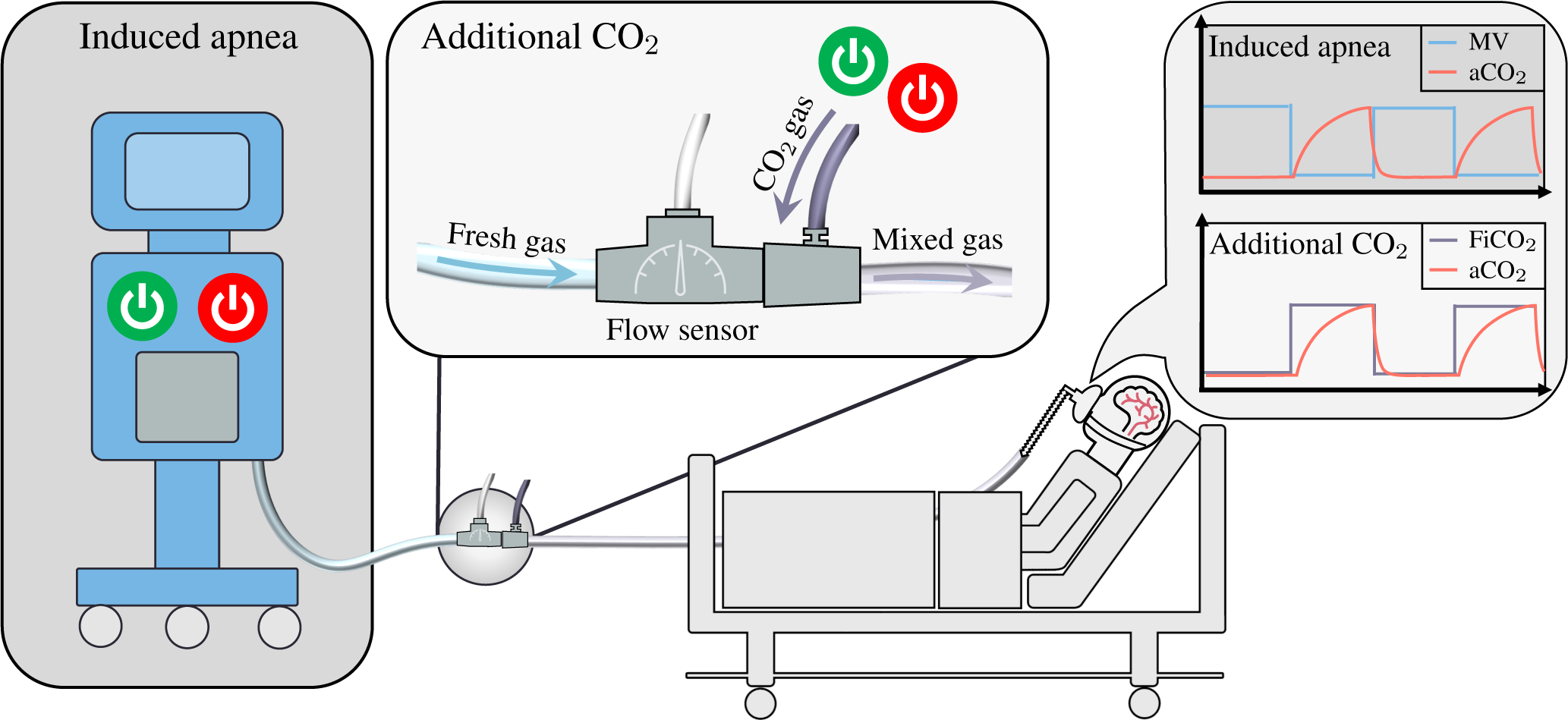
Illustrating the fundamental distinction between two methods employed for the measurement of Cerebrovascular Reactivity in mechanically ventilated patients: Induced Apnea and Additional CO_2_. The Induced Apnea method, commonly referred to as “breath-hold”, produces a hypercapnic stimulus by temporary switching off the ventilator, resulting in a transient cessation of the patient’s minute ventilation (MV). This leads to an increase in arterial CO_2_ levels, which subsequently revert to baseline upon reactivation of the ventilator. This method has been illustrated by Fierstra et al. (2017). In contrast, the Additional CO_2_ method maintains continuous ventilation as the ventilator operates without interruption. Instead, it introduces high-concentration CO_2_ intermittently into the breathing circuit, modulating the composition of inspired gases. The flow of fresh gas is continuously measured through a flow sensor, while a mass-flow controller (not shown) regulates the admixture of CO_2_ to maintain a predetermined target CO_2_ concentration in the inspired gas. Given the uninterrupted operation of the ventilator, continuous monitoring of the patient’s O_2_ and CO_2_ levels ensues, offering an added layer of safety and control.

## 2 MATERIALS AND EQUIPMENT

In this section, we outline the material and equipment employed in evaluating the Additional CO_2_ method, directing readers to the supplementary material for a comprehensive description of specific components used.

### 2.1 Additional CO_2_ System

A prototype system, Additional CO_2_ System, was devised to assess the Additional CO_2_ method, consisting of four primary components: gas source (100 % CO_2_, 5 L canister, AirLiquide), a control unit (including a microcontroller, Arduino Beetle, DFRobotic), gas control (flow sensor, SFM3200, Sensirion and mass-flow controller, SFC5400, Sensirion) and graphical user-interface (GUI, Python program running on a laptop, in-house developed). The flow sensor was read by the control unit, which also managed the mass-flow controller. The proportional relationship between the setpoint of the mass-flow controller and the flow of the flow sensor was computed at the GUI and transmitted to the control unit. The underlying calculation involved the solving of a mass balance equation for a target fractional concentration of inspired CO_2_ (FiCO_2_):

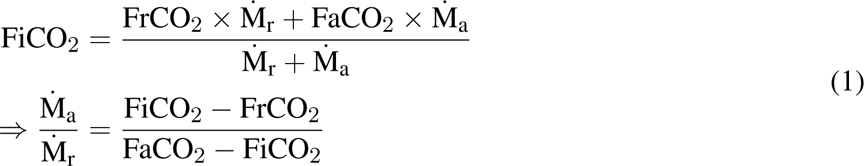

where M_r/a_CO_2_ and F_r/a_CO_2_ are the mass-flow and CO_2_ concentration of the respiratory (r) and additional (a) gas. A consequence of introducing additional CO_2_ in this manner is the concurrent reduction of oxygen concentration in the inspired gas. The user interface also displayed this change in inspired O_2_ levels (FiO_2_) to the user. Moreover, to ensure safety, strict limits were imposed on the maximal and minimal FiCO_2_ and FiO_2_ concentrations, set at 5 % and 19 %, respectively.

To specify the target FiCO_2_ concentration, a user loaded a JSON protocol file containing target values and corresponding time durations via the GUI. For a more comprehensive description of the constructed system and the components employed, refer to section 1.1 in the supplementary material.

### 2.2 Reservoir CO_2_ System

A reference system, modeled after the design by Tancredi et al. (2014), was assembled to facilitate a comparative analysis with our Additional CO_2_ System. This system, from here on referred to as Reservoir CO_2_ System, was established using three mass-flow controllers (SLA5850, Brooks Instrument) connected to sources of oxygen, carbon dioxide and nitrogen. By altering the setpoints of each controller, a specific gas mixture was created and stored in a reservoir, from which a subject would draw breath. The same GUI mentioned earlier was employed to oversee the mass-flow controllers. Users could specify target FiO_2_ and total flow rate, in addition to FiCO_2_ concentrations, utilizing a protocol file similar to the one used for the Additional CO_2_ System. The same constraints on maximal and minimal FiCO_2_ and FiO_2_ concentrations, as described above, remained in effect. For additional information regarding the system and components employed, please consult section 1.2 in the supplementary material.

### 2.3 Ventilator and Test-Lung

To evaluate the Additional CO_2_ System in conjunction with mechanical ventilation, an Anesthesia Workstation (Primus Infinity Empowered, Dräger Medical) was used together with a test-lung (AccuLung, Fluke Biomedical). The Workstation was also used for sampling inspired and expired O_2_ and CO_2_.

### 2.4 Breathing Circuits

Two distinct breathing circuits were employed: one for mechanical ventilations of a test-lung (Ventilator Setup) and another for spontaneous breathing among healthy volunteers (Subject Setup), as illustrated in figure 2. In the Ventilator Setup, which was only used together with the Additional CO_2_ System, the flow sensor was connected to the ventilator’s outlet, followed by a connector with a gas inlet to which the mass-flow controller’s outlet was attached. An empty humidifier was positioned immediately after the connector to serve as a small volume ensuring a uniform gas mixture. A coaxial ventilator tube was affixed to the outlet of the humidifier, and an elbow connector with a sampling port connected the tube to the test-lung.

**Figure 2.**
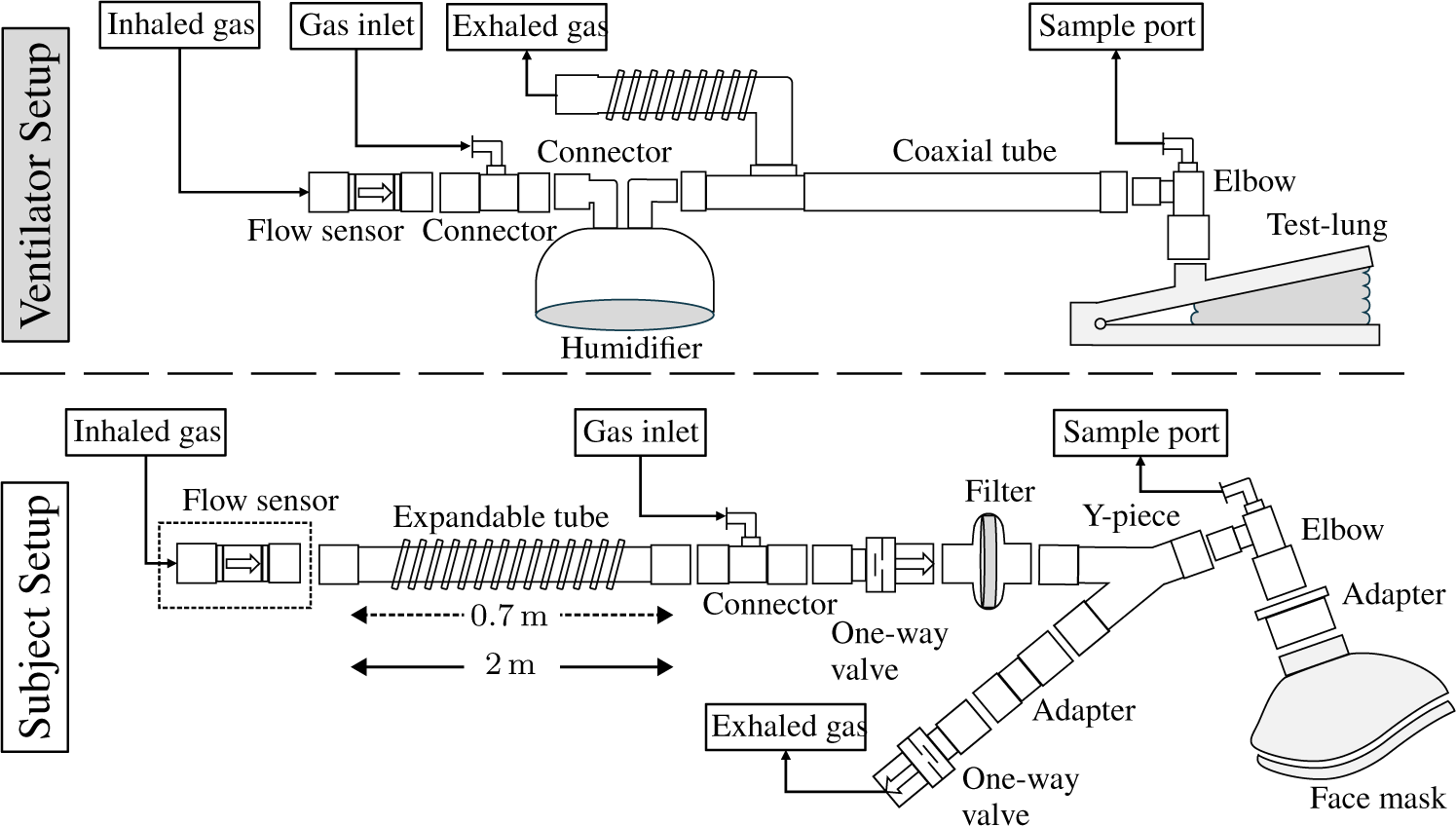
A schematic representation of the respiratory circuits employed in mechanical ventilation of a test-lung (Ventilator Setup) and spontaneous breathing among healthy subjects (Subject Setup). In the Ventilator Setup, the respiratory circuit encompasses a flow sensor affixed to the outlet of the ventilator (not depicted), followed by a connector equipped with a luer-port to facilitate the introduction of additional CO_2_. To ensure the homogeneity of the gas mixture, an empty humidifier was incorporated to enable air mixing. A coaxial tube connected to the humidifier and to the ventilator’s inlet, with the distal end attached to the test-lung via an elbow featuring a sampling port. In the Subject Setup, the configuration of the respiratory circuit differed slightly for the Additional CO_2_ and Reservoir CO_2_ Systems. In the Reservoir CO_2_ configuration, the deployment of a flow sensor was omitted, and the extendable tube was elongated from its minimal length of 0.7 m, as utilized in the Additional CO_2_ configuration, to a length of 2 m, serving as a reservoir for the added gas. The expandable tube was affixed to the gas inlet connector, followed by one-way valves and a Y-connector separating the inhalation and exhalation part of the circuit. A filter prevented particles reaching the subjects who breathing in the circuit through a face mask which was fitted to the head with the help of an adjustable harness (not shown).

In the Subject Setup, the lower part of figure 2 depicts the circuit used. The sole differences between the Additional CO_2_ and Reservoir CO_2_ configurations were the inclusion of the flow sensor (used solely in the Additional CO_2_ System) and the length of the expandable 22 mm tube. In the Reservoir CO_2_ System, the expandable tube functioned as the gas reservoir, as elucidated by Tancredi et al. (2014), and was extended to a length of 2 m, creating a reservoir with a size of 760 mL. Given that normal tidal volumes in adults are approximately 500 mL, this size was deemed sufficient (Hallett et al., 2023). Conversely, for the Additional CO_2_ System, the expandable tube was minimized to 0.7 m. The reason for not completely removing the tube was the desire to maintain the flow sensor away from the center of the MRI scanner to avoid interference when using the system in a full BOLD-CVR setup (see BOLD-CVR Examination section below). Apart from these variances, the breathing circuit remained uniform for both the Additional CO_2_ and Reservoir CO_2_ Systems and comprised a connector with a gas inlet for the addition of pure CO_2_ gas (in the Additional CO_2_ System) or a gas mixture of O_2_, CO_2_, and N_2_ (in the Reservoir CO_2_ System). The direction of gas flow was regulated by two one-way valves, and a filter was added to eliminate particles from the inspired gas. A Y-piece separated the inspiration and expiration segments of the circuit, with an elbow connector featuring a sampling port connecting the Y-piece to the face mask (Mask 7450 V2, Vyaire). An in-house 3D printed adapter was used to accommodate the 22 mm elbow to the 30 mm port of the face mask. For a comprehensive inventory of components used, please refer to table 1.3 in the supplementary material.

## 3 METHOD

### 3.1 Assessment of Inspired CO_2_ Target Accuracy

The primary objective of this study was to assess the accuracy of the proposed Additional CO_2_ method in achieving the desired CO_2_ target levels within the inspired gas. This assessment was conducted under two distinct scenarios: mechanical ventilation and spontaneous breathing.

The FiCO_2_ target function employed in this evaluation encompassed a range of stimulus types, as illustrated in figure 3. These stimuli included three box-stimulus at 1 %, 3 % and 5 % CO_2_, each lasting for 45 s, with an initial 60 s baseline period and a 45 s intermediate baseline. Subsequently, a ramp function was applied, increasing CO_2_ concentration from 0 % to 5 % over 60 s, followed by the first half of a sinusoidal waveform with a peak concentration of 5 % and a time period of 120 s. Finally, a 60 s baseline was appended, resulting in a total protocol duration of approximately 9 min.

**Figure 3.**
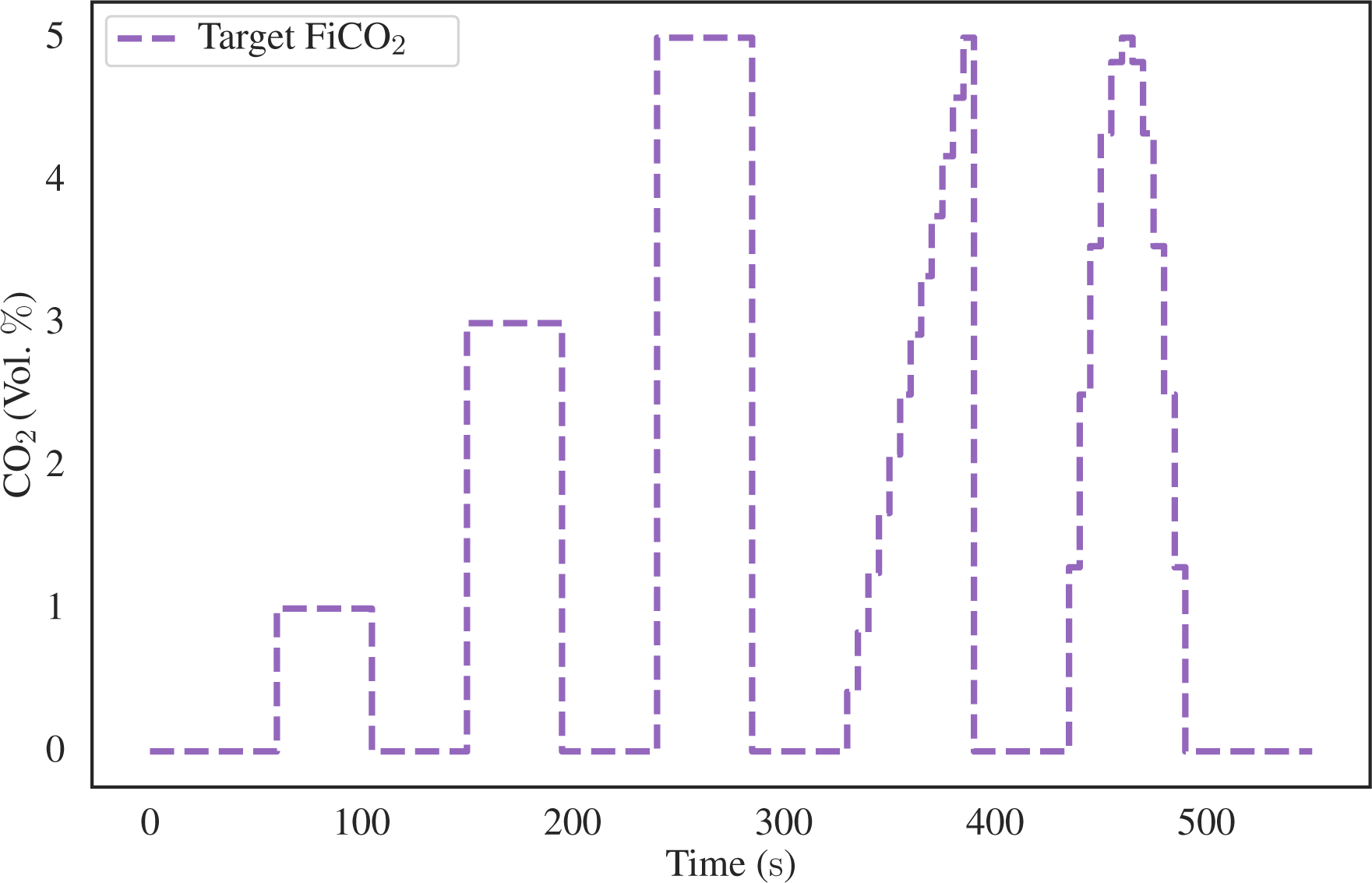
The CO_2_ target function employed for the assessment of the precision of the proposed Additional CO_2_ method. The target function comprises three 45 s box-stimulus intervals, each at distinct CO_2_ concentrations of 1 %, 3 % and 5 %. These stimuli were subsequently followed by a 60 s ramp and half-sinusoidal waveform, both characterized by a peak concentration of 5 % CO_2_. A 45 s baseline was inserted between each stimulus and an initial and final baseline of 60 s duration was also included.

#### 3.1.1 Inspired CO**_2_** Target Accuracy in Ventilator Setup

The accuracy of inspired CO_2_ levels during mechanical ventilation was evaluated using a Primus Anesthesia Workstation in conjunction with an AccuLung test-lung, following the equipment setup delineated in the Materials and Equipment section. To ensure a comprehensive assessment, a variety of ventilator conditions were considered, aligning with the specifications established by the European standard ISO 80601-2-12:2020. Within this standard, two specific categories were explored: volume-control inflation (table 201.104) and pressure-control inflation (table 201.105). Due to limitations in the available settings of the AccuLung test-lung, only the initial seven test cases from each table, totaling 14 test cases, were feasible. The complete list of these test cases is provided in table S4 within the supplementary material.

For each test case (randomized in order), the FiCO_2_ target depicted in figure 3 was administered by the Additional CO_2_ System through the breathing circuit shown in the upper portion of figure 2. The Primus Workstation continuously sampled both oxygen and carbon dioxide at an approximate frequency of 60 Hz. Utilizing the sampled O_2_ and CO_2_ curves, the inspired O_2_ and CO_2_ levels were calculated by an automated Python script. The script identified the inspiratory phase and computed both the peak and baseline levels of O_2_ and CO_2_ to measure the variability within each inspiration. These values were subsequently interpolated to ensure uniform sampling across all 14 ventilator test cases which enabled aggregation and computation of mean plus confidence intervals using the built-in functionalities of the Seaborn package in Python.

To effectively compare the aggregated values with the target FiO_2_ and FiCO_2_ levels, the aggregated data was time shifted 8 s to compensate for the sampling delay of 3 s and the presence of dead space within the ventilator tubing. This dead space necessitated multiple breaths before any alteration in inspired CO_2_ concentrations would manifest at the sampling port. While it is acknowledged that individual test runs would have experienced distinct time delays, accounting for variations in tidal volume and respiratory rate, it was determined that the uniform application of the same delay to all runs introduced a relatively minor error when compared with other sources of variation, such as the temporal misalignment between the onset of the stimulus and the start of the subsequent breath.

#### 3.1.2 Inspired CO_2_ Target Accuracy in Subject Setup

To conduct a comprehensive evaluation of the Additional CO_2_ method, six healthy subjects (aged between 25 and 42, 3 males and 3 females) were recruited to assess accuracy of inspired CO_2_ in spontaneous breathing. Additionally, we made a comparative analysis between our proposed system: Additional CO_2_ System, and the previously described system outlined by Tancredi et al. (2014): Reservoir CO_2_ System.

The recruitment process strictly adhered to the principles outlined in the Helsinki Declaration, and ethical approval was obtained from the Swedish Ethical Review Authority (reference number: 2021-04825). Prior to their participation, the selected subjects underwent a screening process to ascertain the absence of pulmonary diseases or other chronic health conditions.

In the Materials and Equipment section, the experimental configurations used for the Additional CO_2_ and Reservoir CO_2_ Systems are described. Various sizes of face masks, ranging from XS to L, were made available and selected based on an optimal fit for each subject’s facial dimensions. These face masks were secured onto the subjects’ faces using harnesses, which allowed for adjustment to ensure an airtight seal. To verify the effectiveness of the seal, subjects were instructed to block the face mask’s inlet and attempt to breathe. If air leakage was detected, adjustments were made until a tight seal was achieved. It is noteworthy that, in some instances, particularly among subjects with facial hair, attaining a complete seal proved challenging, and, in a few cases, it remained unattainable. Importantly, this limitation was consistent across both the Additional CO_2_ and Reservoir CO_2_ configurations and was thus accepted as an inherent limitation to both methods.

The acquisition of O_2_ and CO_2_ concentrations was made by the Primus ventilator, now operating in surveillance mode. Notably, the ventilator was not linked to the inspiration and expiration portions of the breathing circuit (lower part of figure 2), as these components remained open to the surrounding room environment.

The same FiCO_2_ target protocol employed in the mechanical ventilation configuration was used (figure 3). Subjects were instructed to maintain calm and normal breathing while the target stimulus was administered. The experiment was repeated for both the Additional CO_2_ and Reservoir CO_2_ configurations for each subject, resulting in a total of 12 experimental runs. The sequence in which these two methods were employed was randomized in blocks to mitigate any order effects. Furthermore, it is relevant to mention that the Reservoir CO_2_ method allowed for the specification of the total flow of fresh gas and inspired O_2_ levels, which is not actively controlled in the Additional CO_2_ method. To facilitate comparison between the two methods, the target FiO_2_ level in the Reservoir CO_2_ System was set to 21 %, approximately corresponding to the ambient room concentration. Regrettably, achieving the 15 L flow rate of fresh gas as proposed by Tancredi et al. (2014), was unattainable due to constraints imposed by the maximum flow capacity of the mass-flow controllers, which was limited to 10 L. Instead, we chose to use 8 L of fresh gas flow, which is approximately the upper limit of common minute ventilation in healthy adults which range between 6 L to 8 L (Hallett et al., 2023; Sapra et al., 2023).

A semi-automated Python script was used to calculate inspired and end-tidal O_2_ and CO_2_ values for each experimental run. This script generated initial estimations for inspired and end-tidal values, which could be further refined by the user as necessary. The need for manual intervention stemmed from the inherent complexity of sampled O_2_ and CO_2_ curves in spontaneous breathing subjects, as these curves exhibited greater variability, multiple peaks, and valleys compared to the more stable curves observed in passive ventilated test-lung settings. To fully capture the range of values within each inspiration, 1-3 data points were tracked. Data aggregation across multiple runs and subjects was facilitated by categorizing the inspired and end-tidal O_2_/CO_2_ values into 5 s bins, a process that also introduced some degree of smoothing. Consequently, the need for time-shifting values to compensate for the sampling delay of 3 s was eliminated. Inspired values from individual runs could then be aggregated across all subjects and directly compared to the target FiO_2_ and FiCO_2_ levels, again with the help of Seaborn package in Python. In the case of end-tidal O_2_ and CO_2_ values, baseline subject variations were first removed by subtracting the mean end-tidal value during the initial 60 s of each run before aggregating across subjects.

### 3.2 BOLD-CVR Examination

In the context of assessing the Additional CO_2_ method as a technique for CVR measurement, it is essential to note that the conventional approach to conducting CVR examination relies on utilizing the BOLD signal as a surrogate measure of blood flow. We therefore evaluated the Additional CO_2_ method in an MRI-environment in a subset of two subjects. They underwent BOLD-CVR examinations using both the Additional CO_2_ and Reservoir CO_2_ Systems, which were repeated twice in a test-retest experimental design, yielding a total of four runs per subject. The same breathing circuit from the target accuracy assessment of inspired CO_2_ (lower part of figure 2) was used. Detailed information regarding the experiment, including MRI and CO_2_ protocols, as well as the generation of CVR maps, can be found in section 2 of the supplementary material.

## 4 RESULTS

### 4.1 Inspired CO_2_ and O_2_ Target Accuracy

The present section delves into the outcomes of the experiment aimed at evaluating the target accuracy of inspired CO_2_. To illustrate the analysis, figure 4 shows example datasets. In the uppermost section of figure 4, the Additional CO_2_ System with the Ventilator Setup is depicted, along with a randomly selected test-case. The middle section showcases the Additional CO_2_ System with the Subject Setup in conjunction with a random subject, while the lowermost part illustrates the Reservoir CO_2_ System with the same subject. In all instances, the same target FiCO_2_ protocol, as detailed in figure 3, was used. Each figure offers insight into the sampled CO_2_ levels and the calculated inspired/end-tidal CO_2_ values. It should be noted that the test-lung does not possess end-tidal CO_2_ values.

**Figure 4.**
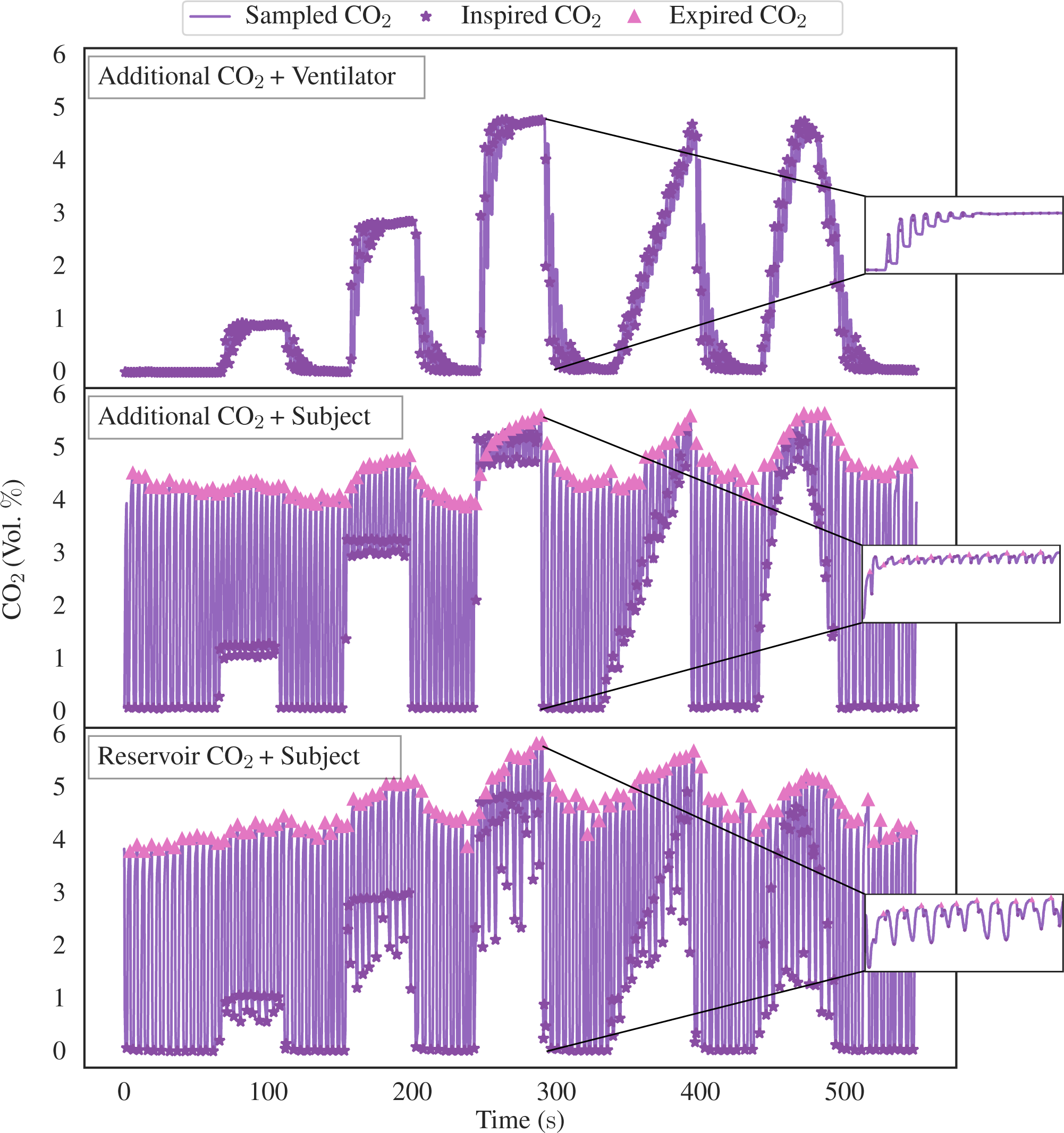
Illustrative data pertaining to FiCO_2_ target accuracy assessment, delineating the three distinct experimental configurations: the Additional CO_2_ System + Ventilator Setup (depicted in the top graph), the Additional CO_2_ System + Subject Setup (displayed in the middle graph), and the Reservoir CO_2_ System + Subject Setup (depicted in the bottom graph). All three instances use the identical target function as showcased in figure 3. Upon closer examination within the magnified window, we see the dynamic fluctuations in CO_2_ concentration throughout the 5 % box-stimulus. It becomes evident that the rise time of CO_2_ in the Ventilator Setup exhibits a substantially slower response in comparison to the other two configurations. Furthermore, the Reservoir CO_2_ System manifests a markedly greater degree of variability in inspired CO_2_ levels when compared with the Additional CO_2_ System.

To ascertain the performance of the experimental configurations the data from all runs were aggregated over all test-cases/subjects as outlined in the Method section. This process has enabled the computation of the mean and a 95 % confidence interval for the inspired/end-tidal CO_2_, as displayed in figure 5. Note that end-tidal values has been converted from volume percentage to partial pressure, assuming an atmospheric pressure of 760 mmHg.

**Figure 5.**
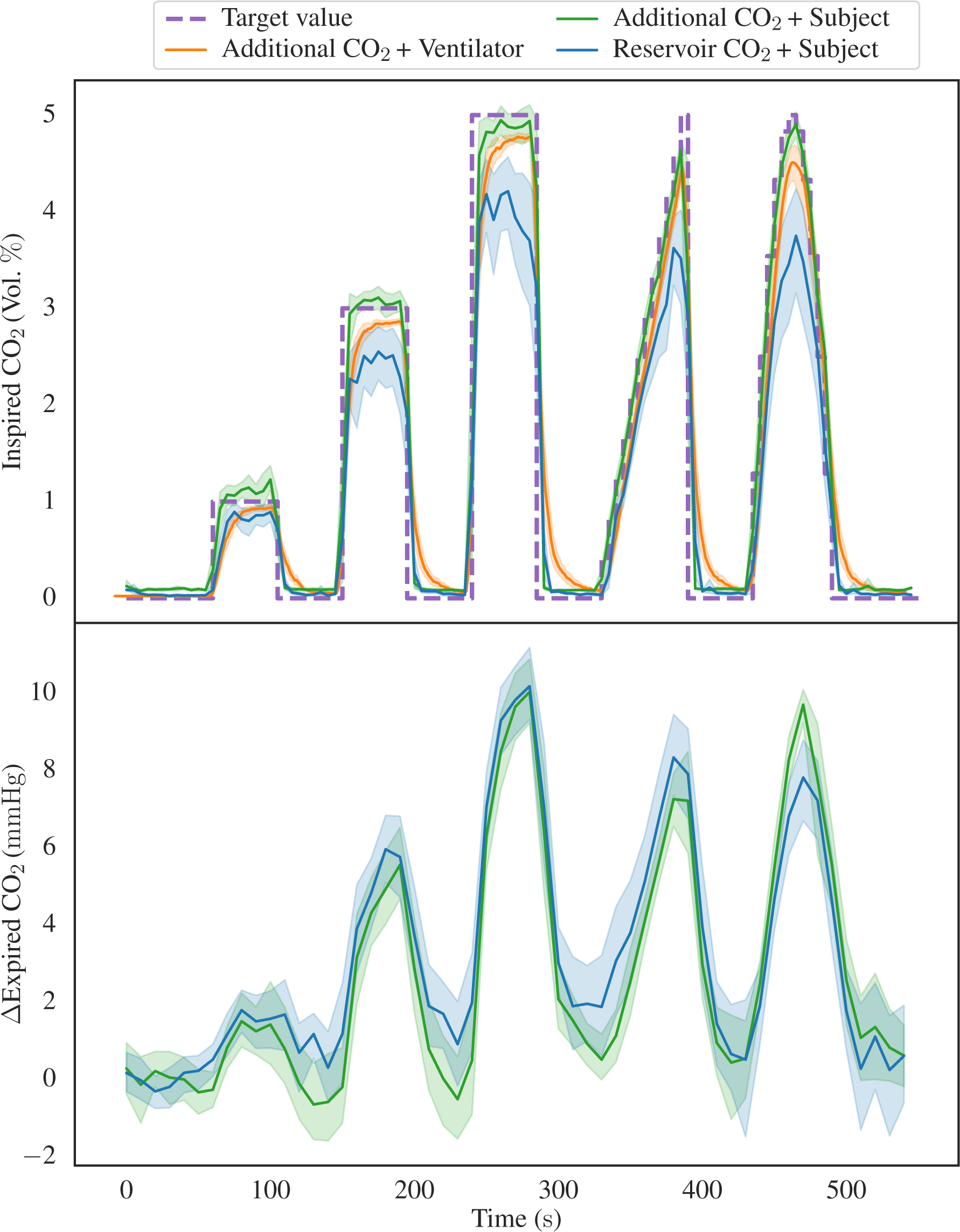
Illustrating the aggregated inspired CO_2_ levels in the top graph with data from the three distinct configurations: Additional CO_2_ System + Ventilator Setup, Additional CO_2_ System + Subject Setup, and Reservoir CO_2_ System + Subject Setup. The data is depicted in terms of both the mean values and a 95 % confidence interval, alongside the target FiCO_2_. Notably, it becomes evident that the Additional CO_2_ System outperforms the Reservoir CO_2_ System in its ability to attain diverse CO_2_ levels, although a consistent undershoot is observed in the Ventilator Setup. In the lower graph, the aggregated end-tidal CO_2_ values for the two sets of subject data are presented. It is worth observing that, despite the visible variability in the inspired CO_2_, disparities in the end-tidal CO_2_ are less obvious.

The accuracy and precision of each setup, assessed by the mean deviation between the target and measured FiCO_2_, was quantified for each type of stimulus. The mean deviations, after eliminating transition periods for the box-stimuli (initial 10 s and final 5 s) and ramp-stimulus (final 5 s), are summarized in figure 6.

**Figure 6.**
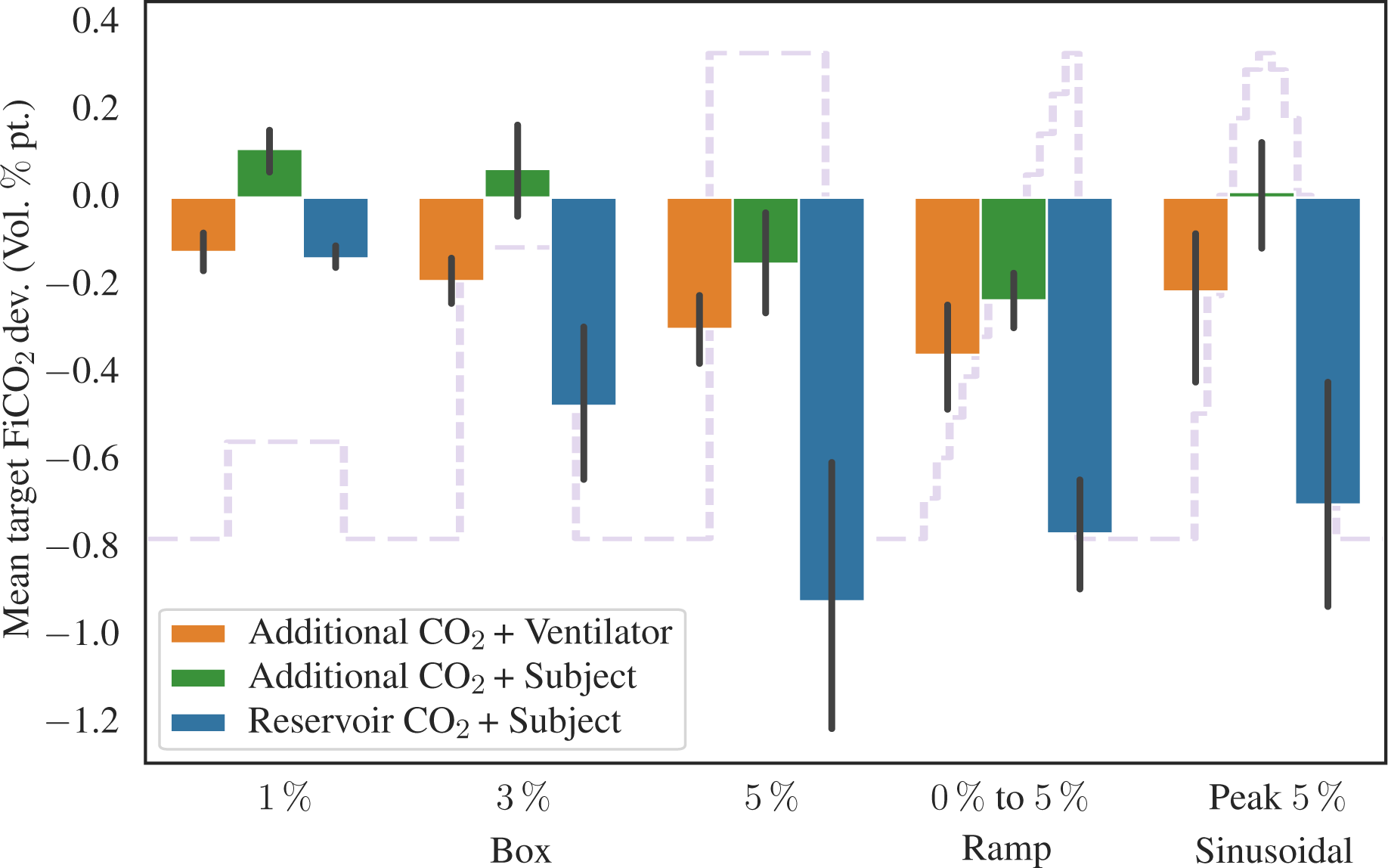
Showing the mean target FiCO_2_ deviation, in volume percentage point, for the different stimuli and experimental configurations. The target function from figure 3 is shown in the background. Transition periods for the box-stimuli (initial 10 s and final 5 s) and ramp-stimulus (final 5 s), have been removed when calculating the mean deviation. Also shown are 95 %-confidence interval error bars.

Next, we redirect our attention toward the aggregated values of inspired and end-tidal oxygen, as depicted in figure 7, with the uppermost graph showing the inspired O_2_ levels, and the lower graph showcasing the end-tidal O_2_ values. We restrict us to presentation of data from the Additional CO_2_ System + Subject Setup and Reservoir CO_2_ System + Subject Setup configurations. Even though both systems target the same baseline O_2_ concentration, 21 %, the Additional CO_2_ System does so passively by the usage of room air, which is not exactly 21 %. To facilitate direct comparison between the two configurations, the measured and target FiO_2_ levels have been normalized by their baseline value in figure 7.

**Figure 7.**
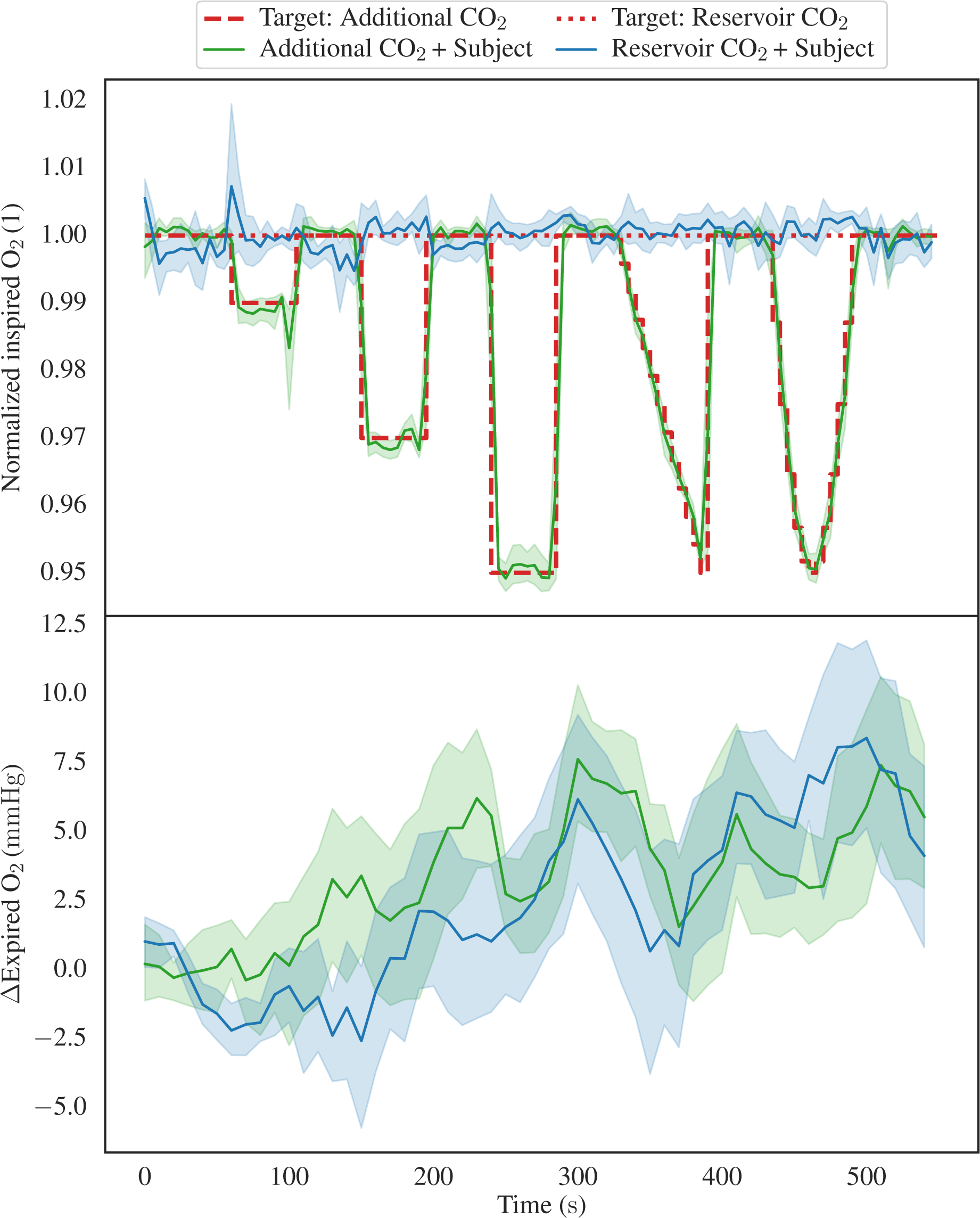
Showing the inspired/end-tidal O_2_ levels, focusing exclusively on the Additional CO_2_ System + Subject Setup and Reservoir CO_2_ System + Subject Setup configurations. The top graph displays the inspired O_2_, including both the mean values and 95 % confidence intervals. It is noteworthy that two distinct target function are depicted, in the Reservoir CO_2_ configuration, the FiO_2_ remains constant, while in the Additional CO_2_ configuration, it varies due to the introduction of additional carbon dioxide. Further, the measured and target FiO_2_ values have been normalized by their baseline value to allow for direct comparison between the two methods. The lower graph presents the aggregated end-tidal O_2_ levels. Notably, both graphs exhibit analogous trends characterized by an increase in O_2_ levels over time, despite the notable disparity in inspired O_2_ concentrations between the two experimental configurations.

### 4.2 BOLD-CVR Experiment

In figure 8, we present illustrative CVR maps obtained through the application of the Additional CO_2_ (A-CO_2_) and Reservoir CO_2_ (R-CO_2_) Systems within a single subject. It is imperative to emphasize that our objective is not to derive quantitative inferences, however, figure 8 does unveil a qualitative congruence in the CVR maps yielded by both methods.

**Figure 8.**
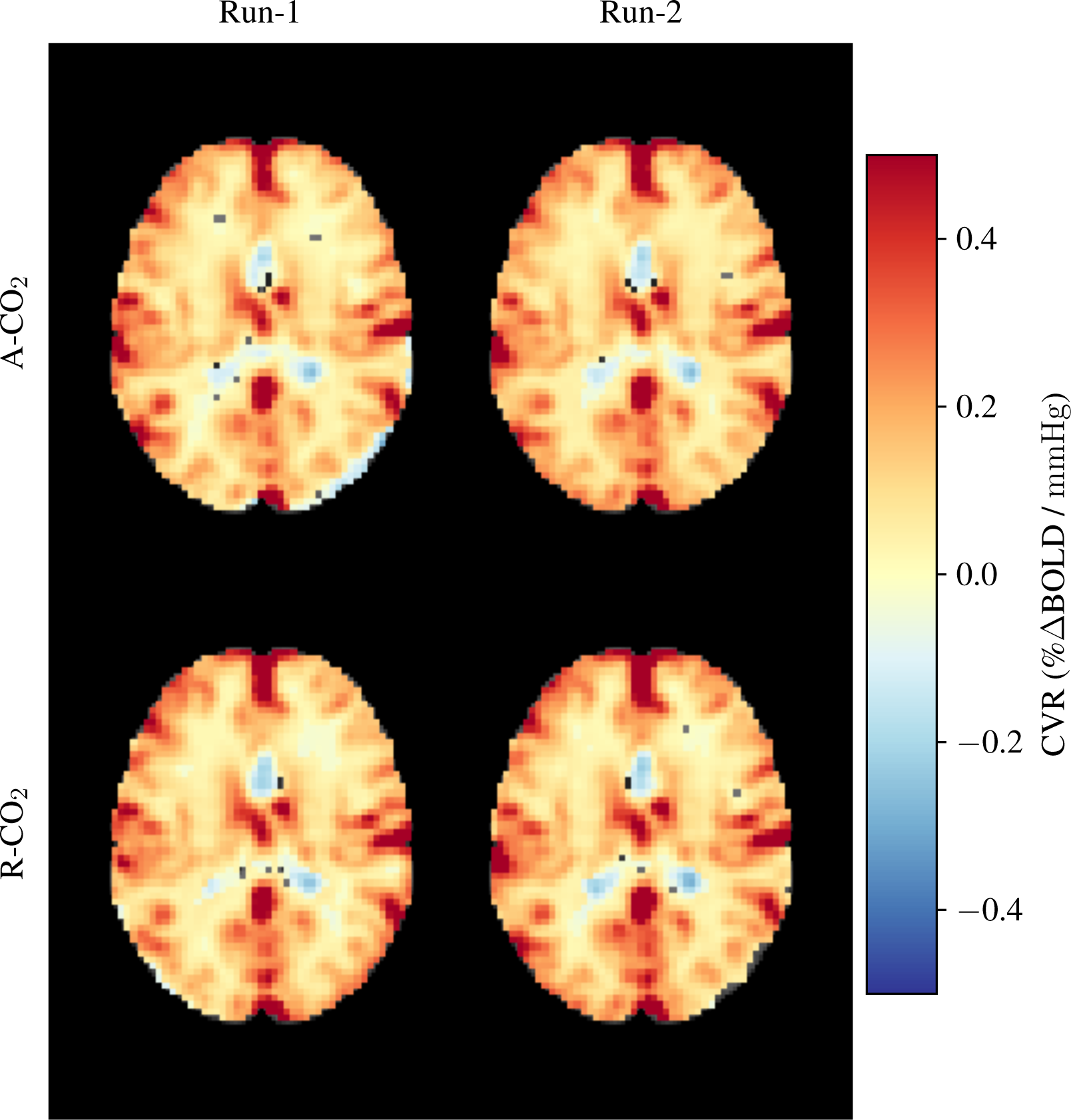
Exemplar CVR maps obtained from a single participant. The upper row showcases CVR maps generated utilizing the Additional CO_2_ System, while the lower row exhibits CVR maps derived from the Reservoir CO_2_ System. These measurements were replicated twice for each system configuration.

## 5 DISCUSSION

### 5.1 Inspired CO_2_ Target Attainment

When examining the illustrative data presented in figure 4, noticeable disparities among the three distinct configurations (Additional CO_2_ System + Ventilator Setup, Additional CO_2_ System + Subject Setup, and Reservoir CO_2_ System + Subject Setup) become evident. First, the ventilator configuration displays slower CO_2_ response compared to the two subject configurations. The distinct behavior arises primarily from the gas inlet’s placement within these setups. In the Ventilator Setup, the inlet is located proximal to the flow sensor, whereas, in the Subject Setup, the inlet is proximal to the sampling port (as depicted in 2). This discrepancy dictates the rate of CO_2_ level alteration due to the volume within the tubes, as air is propelled forward in fixed tidal increments. The rationale for not placing the gas inlet proximal to the sampling port in the Ventilator Setup is the need to minimize the distance between the flow sensor and the gas inlet due to the overpressure, characteristic of ventilator operation. Otherwise, a substantial disparity arises between the flow sensor’s measured flow and the gas delivered by the mass-flow controller. Secondarily, the Additional CO_2_ configuration for the selected subject displays considerably less variance in inspired CO_2_ values in comparison to the Reservoir CO_2_ configuration. A closer examination of the data reveals that in the Reservoir CO_2_ configuration, the initial inspired CO_2_ closely approximates the target value but then suddenly declines toward zero. This behavior indicates that gas within the reservoir has not undergone a complete exchange, implying that the influx of fresh gas is inadequate. This limitation appears reasonable, given that only 8 L of fresh gas is used in contrast to the 15 L used in the study by Tancredi et al. (2014), see section Inspired CO_2_ Target Accuracy in Subject Setup. This should be acknowledged as a constraint in our experimental system. Nevertheless, it underscores the imperative need for a substantially greater flow rate than one would expect by merely considering minute ventilation, which typically ranges between 6 L to 8 L in healthy adults (Hallett et al., 2023; Sapra et al., 2023).

Directing our focus to the inspired CO_2_ levels (upper portion of figure 5), it becomes apparent that the Additional CO_2_ method consistently adheres to the target value within the subject dataset. It notably outperforms the Reservoir CO_2_ method, a distinction further quantified in figure 6, which elucidates the mean divergence between target and measured FiCO_2_. However, as previously mentioned, this disparity likely arises from an insufficient flow of fresh gas in the Reservoir CO_2_ System. Indeed, the end-tidal CO_2_ levels (lower portion of figure 5), which reflect alveolar concentration, exhibit small discrepancy between the two methods. This outcome is not surprising since it is primarily the initial inspired gas portion that reaches the alveoli, with subsequent inspired gas lingering in the anatomical dead space of the conducting airways. Therefore, the constraint associated with limited fresh gas flow of the Reservoir CO_2_ method may not be significant if the flow is sufficient to cover alveolar ventilation.

In revisiting the upper portion of figure 5, it is noteworthy that the Additional CO_2_ System consistently undershoots the target value in the Ventilator Setup. While the offset is relatively small, amounting to less than 0.4 percentage points (see figure 6), understanding the rationale behind this deviation holds intrinsic value. One plausible explanation pertains to the sensitivity of the SFM3200 flow sensor to laminar flow. To ensure accurate measurements, the manufacturer, Sensirion, underscores the necessity of establishing laminar flow. Preliminary assessments suggest that turbulent flow yields higher readings than laminar flow. Consequently, if the ventilator produces a higher proportion of laminar gas flow relative to the gas used during the calibration of the Additional CO_2_ System (incorporating the flow sensor and mass-flow controller), this might elucidate the observed persistent undershoot evident in figure 5. However, further investigations are requisite to explain this apparent discrepancy in the Ventilator Setup. Although any offset is undesirable from a standpoint of repeatability, a consistent target undershoot arguably fares better than a consistent target overshoot concerning subject safety and tolerance.

### 5.2 Oxygen Concentration Outcome

Figure 7 shows the inspired and end-tidal O_2_ concentrations for the two subject configurations, using the Additional CO_2_ and Reservoir CO_2_ Systems. As delineated in the Method section, the Additional CO_2_ method does not actively regulate O_2_ concentration, rather, it manifests as a direct consequence of adding CO_2_ to the inspired gas. Hence, it is unsurprising that the target FiO_2_ level (depicted as the dashed red line in the upper portion of figure 7) inversely mirrors the target FiCO_2_ level. In the Reservoir CO_2_ method, O_2_ levels are actively controlled by the system and have been maintained at a constant 21 % (as indicated by the dotted red line in figure 7). To facilitate a comparison between the two methods, the measured and target FiO_2_ values have been scaled by their baseline value. Given the conspicuous dissimilarities in measured FiO_2_ levels, one might reasonably anticipate notable discrepancies in end-tidal O_2_ levels. However, a close examination of the lower segment of figure 7 reveals a lack of pronounced differentiation between the two methods. This phenomenon arises from the recognition that inspired concentration is not the sole determinant of end-tidal values. Variations in minute ventilation, by increased or decreased breath frequency and depth, typically lead to concurrent change in end-tidal O_2_ (and CO_2_) values. Inspecting the end-tidal O_2_ curve for the Reservoir CO_2_ dataset reveals a progressive elevation over time, signifying an increasing minute ventilation as the experiment unfolds, even though the inspired O_2_ concentration stays fixed, a phenomenon attributed to the automatic triggering of reflexes to stimulate deeper and more frequent breaths when CO_2_ is inspired (Carr et al., 2021). Similarly, in the Additional CO_2_ configuration, end-tidal O_2_ levels appear to rise as the experiment progresses. Hence, although the inspired O_2_ levels exhibit fluctuation in the Additional CO_2_ method, the effect is obscured by variations in minute ventilation, thus attenuating the disparities between the Additional CO_2_ and Reservoir CO_2_ methods. Advanced control systems, such as prospective end-tidal targeting, account for these changes in minute ventilation to provide a more precise and reproducible stimulus (Slessarev et al., 2007).

It is worthy of note that in the Ventilator Setup, tidal volumes, and consequently minute ventilation, remained constant when the test-lung was ventilated using pressure-control inflation, but not when volume-control was employed. This discrepancy is understandable since, in volume-control ventilation, the ventilator administers a predefined tidal volume, with any additional CO_2_ gas adding to this volume. Conversely, pressure-control ventilation involves the establishment of a fixed inspiration pressure (P_insp_) at the outset of each breath, maintained for a predetermined duration (T_insp_). In such scenarios, tidal volume becomes dependent solely upon P_insp_, T_insp_ and the compliance of the test-lung (or patient), why the addition of CO_2_ gas does not alter the tidal volume. Therefore, in real-life mechanical ventilation of a patient, a reduction in end-tidal O_2_ levels is to be anticipated when employing the Additional CO_2_ method to administer CO_2_.

### 5.3 BOLD-CVR Outcome

We examined the BOLD-CVR in two research subjects. The dataset depicted in figure 8 presents initial findings, serving as an illustrative demonstration of the feasibility of our proposed Additional CO_2_ method. It is crucial to underscore, nonetheless, that a more extensive, in-depth inquiry is imperative to assess the applicability of the Additional CO_2_ method in an MRI context. For a more detailed exploration of the BOLD-CVR experiment, we direct interested readers to section 2 in the supplementary materials.

### 5.4 Limitations

In evaluating the Additional CO_2_ method in mechanical ventilation, we only used a test-lung, with no human subject being ventilated. Further, our investigation only considered two types of ventilation mode: volume-control and pressure-control.

## 6 SUMMARY

The contemporary landscape of CVR research predominantly features investigations conducted in spontaneous breathing subjects, with limited attention directed towards individuals undergoing mechanical ventilation. A notable constraint contributing to this disparity resides in the lack of suitable apparatus for executing CO_2_ gas challenges within a ventilator-dependent setting. Consequently, CVR assessments in ventilated patients have conventionally resorted to alternative stimuli, such as induced apnea (breath-hold), entailing cyclic activation and deactivation of the ventilator.

In the present work, we propose a new method, which collaboratively interfaces with mechanical ventilation to administer a variable amount of CO_2_, referred to as Additional CO_2_. We systematically assess the precision of our proposed method in regulating the inspired CO_2_ levels and compare its performance against an established method that relies on a gas reservoir containing a mixture of CO_2_ at varying concentrations. Furthermore, we evaluate the compatibility of our devised system within an MRI environment, conducting a BOLD-CVR study.

Our findings support the efficacy of our method in maintaining precise inspired CO_2_ levels in both mechanically ventilation and in spontaneous breathing. Moreover, it can integrate with an MRI scanner to generate BOLD-CVR maps. We hope that these results will facilitate future research of CVR examinations among mechanical ventilated patients in the near future.

## AUTHOR CONTRIBUTIONS

Gustav Magnusson conducted the experimental research that underpins this article, under the mentorship and guidance of the remaining co-authors. The collective contribution of all authors involved the critical review and incorporation of revisions into the ultimate manuscript, which was principally authored by Gustav Magnusson.

## Data Availability

All data produced in the present study are available upon reasonable request to the authors

## ACKNOWLEDGMENTS

Acknowledgments are extended to the Center for Magnetic Resonance Research (CMRR), University of Minnesota, USA, for providing the Multi-Band Multi-Echo BOLD sequence used in this study.

