## Supplementary Material for "High Inspired CO_2_ Target Accuracy in Mechanical Ventilation and Spontaneous Breathing Using the Additional CO_2_ Method"

#### 1 SYSTEM DESCRIPTIONS AND COMPONENTS

Figure S1 illustrates the configuration of the Additional $CO_2$ System (on the left) and the Reservoir $CO_2$ System (on the right) within a magnetic resonance imaging (MRI) environment. In the Additional $CO_2$ configuration, certain integral components, namely the flow sensor, mass-flow controller, and control unit, are positioned within the confines of the MRI scanner room. In the Reservoir $CO_2$ configuration, all associated components are situated external to the MRI scanner room.

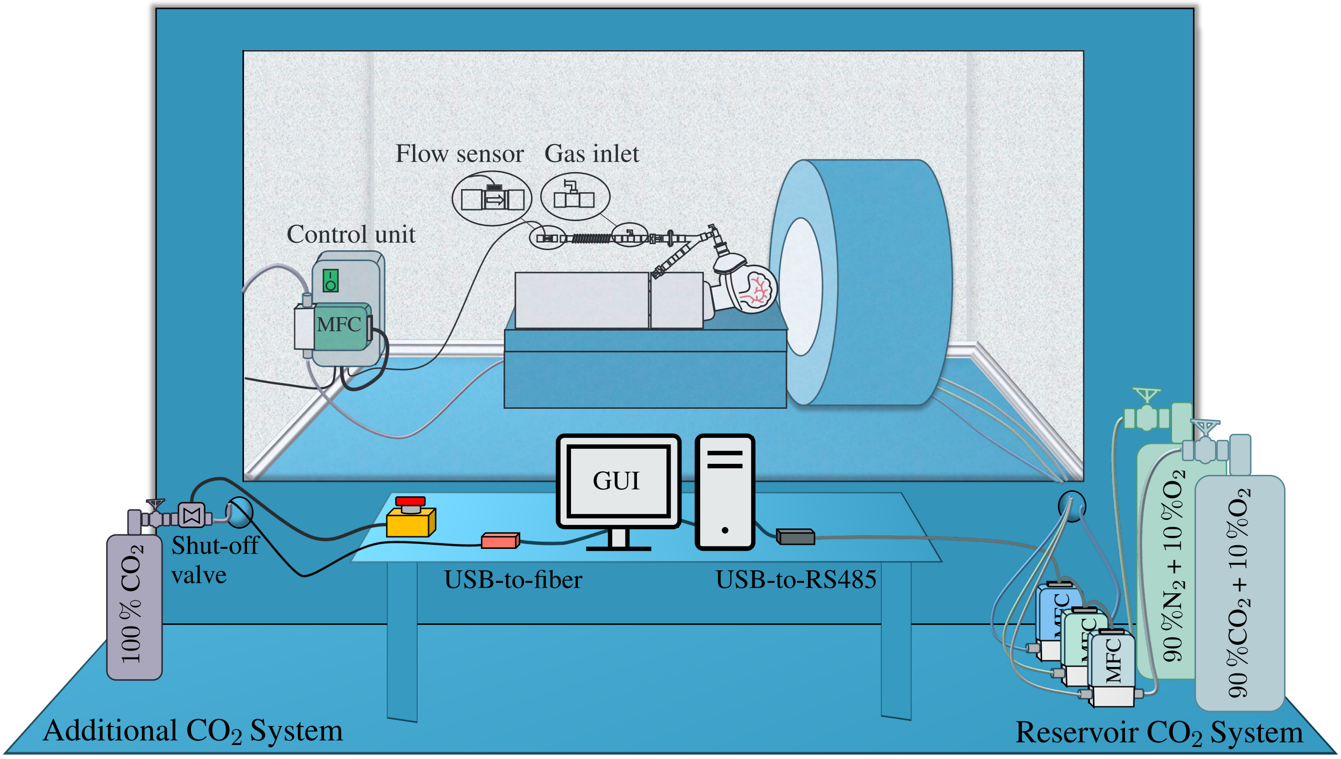


**Figure S1.** Illustrating the Additional $CO_2$ and Reservoir $CO_2$ Systems within an MRI environment. In the Additional $CO_2$ configuration, the flow sensor, mass-flow controller, and control unit are situated within the confines of the MRI scanner room. This placement choice is required by the necessity to maintain a minimal $CO_2$ tube length (less than $5\,\mathrm{m}$) between the mass-flow controller and the inlet in the breathing circuit to ensure precise targeting, see primary article figure 2. The mass-flow controller and control unit are affixed to the wall, while the flow sensor is affixed to the breathing circuit, which is positioned on the scanner board. It is worth noting that the flow sensor remains external to the MRI scanner opening, its position adjusted by the expandable tube in the breathing circuit. A computer-based graphical user interface (GUI) and a $100\,\%$ $CO_2$ gas cylinder are both positioned outside the scanner room, connected via tubing and optical cables that traverse through wave guides into the scanner room. Furthermore, the figure illustrates the presence of an emergency shut-off valve, which provides the means for rapid termination of the $CO_2$ supply. In the Reservoir $CO_2$ configuration, conversely, all equipment is situated external to the scanner room, with only the gas tubes extending into the room through wave guides to establish connections between the mass-flow controllers and the breathing circuit. The expandable tube in this configuration is extended to a length of $2\,\mathrm{m}$, serving as a reservoir. Notably, the flow sensor is not utilized in the Reservoir $CO_2$ setup, nor is it affixed to the breathing circuit.

##### 1.1 Additional $CO_2$ System

The Additional $CO_2$ System is made of various components, which may be categorized into the following main components: $CO_2$ gas supply, Gas control, Control unit, and the Graphical User Interface (GUI). These components, along with their respective subcomponents, are detailed in table 1.1 and are visually depicted in the left portion of figure S1.

The $CO_2$ gas supply is made of a gas cylinder with $100\,\%$ pure $CO_2$, affixed to a shut-off valve, thereby facilitating rapid cessation of gas flow to the mass-flow controller. The Control unit, housing a microcontroller and power supply, controls the setpoint of the mass-flow controller via RS485 protocol and the read the flow sensor via I2C communication to enable a proportional gas flow of $CO_2$ into the primary respiratory airflow. The required proportional gas flow is calculated at the GUI, given a desired target Fraction of Inspired Carbon Dioxide ($FiCO_2$), and subsequently communicated to the Control unit through a fiber optic link.

##### 1.2 Reservoir $CO_2$ System

The Reservoir $CO_2$ System comprises three fundamental components: a $CO_2$/$N_2$/$O_2$ gas supply unit, a gas control system, and a GUI. These constituent elements, along with their respective subcomponents, are detailed in table 1.2 and depicted on the right-hand side of figure S1. The three gas supplies contain $90\,\%$ $CO_2$ + $10\,\%$ $O_2$, $90\,\%$ $N_2$ + $10\,\%$ $O_2$ and $100\,\%$ $O_2$, respectively, and are connected to mass-flow controller to enable precise formulation of gas mixtures. The setpoint for each mass-flow controller is calculated at the GUI, given a target concentration and overall gas flow, and communicated over RS485. The outlets of the mass-flow controllers are attached to tubes which and combined using a 4-way connector before being attached to the gas inlet of breathing circuit, see figure 2 in the primary article.

##### 1.3 Breathing Circuit

The constituents of the two respiratory circuits employed, depicted in figure 2 of the primary article, are listed in table 1.3.

#### 2 BOLD-CVR EXPERIMENT

The final phase of our investigation entailed a test-retest examination of Blood Oxygenation Level Dependent Cerebrovascular Reactivity (BOLD-CVR) with the aim of making a qualitative comparison between the Additional $CO_2$ and Reservoir $CO_2$ methods. The experimental configuration is depicted in figure S1.

##### 2.1 MRI Protocol

A $3\,\mathrm{T}$ Siemens Prisma Magnetic Resonance scanner was employed to monitor alterations in cerebral blood flow resulting from the inhalation of carbon dioxide by the subjects. The MRI protocol encompassed the acquisition of an anatomical T1-weighted MPRAGE scan for image registration. Parameters for this scan were as follows: flip-angle = $8°$, TR = $2.3\,\mathrm{s}$, TE = $2.36\,\mathrm{ms}$, TI = $0.9\,\mathrm{s}$, GRAPPA = 3, reference lines = 24, voxel-size = $0.9\,\mathrm{mm} \times 0.87\,\mathrm{mm} \times 0.87\,\mathrm{mm}$, and matrix = $208 \times 288 \times 288$. For each CVR run, utilizing either the Additional $CO_2$ or Reservoir $CO_2$ technique, a $T2^*$-weighted multi-band (MB) gradient-echo planar imaging sequence (EPI), as provided by the Centre for Magnetic Resonance Research (CMRR, University of Minnesota, USA) was collected (Moeller et al., 2010; Feinberg et al., 2010; Xu et al., 2013)). The parameters for this sequence were: flip-angle = $60°$, TR = $1\,\mathrm{s}$, TE = $12.60\,\mathrm{mm}$, $29.88\,\mathrm{mm}$, $47.14\,\mathrm{mm}$

**Table S1.** Products and manufacturers of components used in the Additional $CO_2$ System.

<table>
<tr><th colspan="2">Component</th><th>Product, Manufacturer</th><th>Comment</th></tr>
<tr><td rowspan="4">$CO_2$ gas supply</td><td>Gas cylinder</td><td>S5 (AL), AirLiquide</td><td>$100\,\%$ $CO_2$, volume $5\,\mathrm{L}$.</td></tr>
<tr><td>Pressure regulator</td><td>Medireg, Linde Gas</td><td>One stage regulator, outlet pressure $4.5\,\mathrm{bar}$.</td></tr>
<tr><td>High pressure tube</td><td>$CO_2$ hospital tube, Linde Gas</td><td>Tube between pressure regulator, shut-off valve and mass-flow controller.</td></tr>
<tr><td>Quick-connectors</td><td>P/N DQSA/DQBA-M-4N-K6-SA, DK-LOK</td><td>Quick connectors with checker valves for easy disconnection of the tube between the shut-off valve and mass-flow controller.</td></tr>
<tr><td rowspan="2">Emergency shut-off</td><td>3/2 way valve</td><td>P/N 126149, Burkert</td><td rowspan="2">The 3/2 way valve is connected to outlet of the pressure regulator, and upon breaking the power supply by pressing the stop button, it closes.</td></tr>
<tr><td>Stop button</td><td>P/N XALK178F, Schneider</td></tr>
<tr><td rowspan="4">Gas control</td><td>Mass-flow controller</td><td>SFC5500, Sensirion</td><td>Calibrated and connected to $CO_2$ gas source. Max. flow: $10\,\mathrm{L\,min^{-1}}$. Communcation via RS485.</td></tr>
<tr><td>Low pressure tube</td><td>Tubclair Al, Tricoflex</td><td>Tube between MFC and gas inlet of breathing circuit. ID: $2\,\mathrm{mm}$, OD: $4\,\mathrm{mm}$.</td></tr>
<tr><td>Flow sensor</td><td>SFM3200, Sensirion</td><td>Flow range: $-100$ to $250\,\mathrm{L\,min^{-1}}$. Communication via I2C.</td></tr>
<tr><td>Quick-connectors</td><td>P/N PMC2404/PMCD1004, Colder</td><td>Quick connectors with checker valve for easy disconnection of the tube connected to the outlet of the mass-flow controller.</td></tr>
<tr><td rowspan="4">Control unit</td><td>Microcontroller</td><td>Arduino Beetle, DFRobot</td><td>Controlling the mass-flow controller and flow sensor and communication with GUI.</td></tr>
<tr><td>Software</td><td>Arduino program, Gustav Magnusson (author)</td><td>In-house developed.</td></tr>
<tr><td>UART-optic-converter</td><td>SPX17510, Sparkfun</td><td>To enable fiber communication between Microcontroller and GUI.</td></tr>
<tr><td>Power supply</td><td>RPT-60, Mean Well</td><td>Power supply for components in Control unit as well as the mass-flow controller and flow sensor</td></tr>
<tr><td rowspan="4">GUI</td><td>Computer</td><td>MacBook Pro, Apple</td><td rowspan="2">Any personal computer with Python installed can be used as computer.</td></tr>
<tr><td>Software</td><td>Python program, Gustav Magnusson (author)</td></tr>
<tr><td>USB-fiber-converter</td><td>SPX17508, Sparkfun</td><td rowspan="2">The USB converter connects to the computer to enable communication with the Control unit over fiber.</td></tr>
<tr><td>Fiber optic cable</td><td>P/N 164-4255, RS PRO</td></tr>
</table>

and $64.42\,\mathrm{mm}$, MB-factor = 4, GRAPPA = 2, reference lines = 24, voxel-size = $3\,\mathrm{mm} \times 3\,\mathrm{mm} \times 3\,\mathrm{mm}$, matrix = $68 \times 68 \times 44$, and #measurement = 345. Additionally, single-band reference (SBRefs) images were saved. Between each CVR run, the subjects was taken out the scanner and asked to sit up before continuing.

##### 2.2 $CO_2$ Protocol

A simplified $CO_2$ stimulation protocol was used, comprising a block-stimulus design with an initial baseline period of $1\,\mathrm{min}$, followed by three alternating cycles of $5\,\%$ and $0\,\%$ $CO_2$ over $45\,\mathrm{s}$ intervals, concluding with an additional minute of baseline recording. To monitor $CO_2$ and $O_2$ levels, a Philips Expression MR400 and its IP5 monitor were employed. The end-tidal $CO_2$ values were made accessible via the RS232 port of IP5 monitor, operating at a data rate of $1\,\mathrm{Hz}$. The recorded end-tidal $CO_2$ values were subsequently utilized for the computation of CVR maps based on the BOLD signal acquisitions.

##### 2.3 Flow Sensor Interference

An unforeseen complication emerged concerning interference of the flow sensor positioned within the MRI scanner. This interference led to disruptions in the communication between the microcontroller and the flow sensor at the onset of scanning. Further investigation revealed that an alteration in the length of the cable connecting the flow sensor to the microcontroller had increased its susceptibility to electromagnetic interference from the MRI scanner. Initial pilot trials employed a shorter $2\,\mathrm{m}$ cable without encountering interference issues, but for subsequent scans, a longer $4.7\,\mathrm{m}$ cable was employed to prevent people tripping over the cable. Unfortunately, this extended cable length closely approximated a multiple of the wavelength of the MRI scanner's RF-field ($2.36\,\mathrm{m}$). By shortening the cable to $4.3\,\mathrm{m}$, most of the disturbances were mitigated, although not eliminated. By carefully securing the cable along the scanner bore, a stable communication link between the microcontroller and the flow sensor was achieved. Two subjects were successfully scanned using this setup. However, when attempting to scan a third subject, the interference issues resurfaced, indicating that the subject's presence also played a role in the interference.

##### 2.4 Preprocessing of MRI Data

The MR data was preprocessed using the automated pipeline fMRIPrep 23.1.4 (Esteban et al., 2019). The T1-weighted images were corrected for intensity non-uniformity and skull-stripped. Volume-based spatial normalization to standard space (MNI152NLin2009cAsym, Fonov et al. (2009)) was performed through nonlinear registration with antsRegistration from the ANTs toolbox (Avants et al., 2008). For each of the BOLD runs a reference volume and its skull-stripped version were generated by using the SBRefs. Head-motion parameters with respect to the BOLD reference (six parameters corresponding three rotation and three translation parameters) was estimated before any spatiotemporal filtering using mcflirt of the FSL toolbox (Jenkinson et al., 2002). BOLD runs were then slice-time corrected to and resampled onto their original, native space by applying the transforms to correct for head-motion. A $T2^*$ was estimated from the preprocessed EPI echoes, by voxel-wise fitting the maximal number of echoes with reliable signal in that voxel to a monoexponential signal decay model with nonlinear regression. The calculated $T2^*$ map was then used to optimally combine preprocessed BOLD across echoes following the method described in Posse et al. (1999). The BOLD reference was then registered to the T1w reference using mri_coreg (FreeSurfer) followed by flirt (FSL) with the boundary-based registration method. The BOLD time-series were finally resampled into standard space (MNI152NLin2009cAsym) utilizing ANTs and the combined BOLD to T1 and T1 to standard space transformation computed earlier.

#### 2.5 Generation of CVR Maps

The generation of CVR maps entailed the utilization of brain-masked BOLD data in conjunction with end-tidal $CO_2$ samples. First, the BOLD data underwent smoothing through the usage of a Gaussian kernel with a full-width-half-maximum of $6\,\mathrm{mm}$. The $CO_2$ data was then temporally adjusted to maximize its correlation with the global BOLD signal, thereby establishing a solid initial alignment. Subsequently, the cross-correlation between the time series of each voxel and end-tidal $CO_2$ was computed, and the $CO_2$ time series was further adjusted on a voxel-by-voxel basis to the maximum correlation. The voxel-specific temporal adjustments were constrained within a range of $\pm 20\,\mathrm{s}$, and the Pearson's correlation p-value, lower-thresholded at 0.05, was used to filter out voxels with low correlation.

The next step involved regressing the voxels against the timeshifted, voxel-specific, end-tidal $CO_2$ data. To account for motion, the previously computed rotation and translation head-motion parameters, including their temporal derivative and power (total of 24 parameters), were included in the regression as confounding timeseries. The resulting end-tidal $CO_2$ regression coefficient was used to calculate the percentage change in BOLD signal per unit change in end-tidal $CO_2$, denoted as $\%\Delta\mathrm{BOLD}/\mathrm{mmHg}$, following the methodology elucidated by Liu et al. (2019).

### 3 VENTILATOR TEST CASES

Fourteen ventilator test cases, prescribed by the European standard ISO 80601-2-12:2020, were incorporated for the purpose of assessing the efficacy and functionality of the Additional $CO_2$ method within the context of mechanical ventilation. These test cases, including test-lung parameters, can be found in table S4.

**Table S2.** Products and manufacturers of components used in the Reservoir $CO_2$ System.

| Component | | Product, Manufacturer | Comment |
|---|---|---|---|
| $CO_2$ gas supply | Gas cylinder | K-sized, AirLiquide | $90\,\%$ $CO_2$ + $10\,\%$ $O_2$, volume $50\,\mathrm{L}$. |
| | Pressure regulator | Multistage, QMT | Two stage regulator, outlet pressure set to $1\,\mathrm{bar}$ |
| | High pressure tube | Super Nobelair, QMT | Tube between pressure regulator and mass-flow controller. |
| $N_2$ gas supply | Gas cylinder | K-sized, AirLiquide | $90\,\%$ $N_2$ + $10\,\%$ $O_2$, volume $50\,\mathrm{L}$. |
| | Pressure regulator | Multistage, QMT | Two stage regulator, outlet pressure set to $1\,\mathrm{bar}$ |
| | High pressure tube | Super Nobelair, QMT | Tube between pressure regulator and mass-flow controller. |
| $O_2$ gas supply | Hospital wall outlet | Unknown | $100\,\%$ $O_2$ |
| | Pressure regulator | Gas outlet regulator, QMT | One stage regulator, outlet pressure set to $1\,\mathrm{bar}$. |
| | High pressure tube | $O_2$ hospital tube, Linde Gas | Tube between pressure regulator and mass-flow controller. |
| Gas control | Mass-flow controllers | SLA5850, Brooks | All three controllers are calibrated and connected to respective gas source. Max. flow: $10\,\mathrm{L\,min^{-1}}$. Communication via RS485. |
| | Low pressure tube | Tygon E3603, Saint-Gobain | Tube between mass-flow controllers and 4-way connecter. ID $6.4\,\mathrm{mm}$, OD $9.5\,\mathrm{mm}$. |
| | 4-way connector | P/N X0-4NK, Colder | Connecting the three mass-flow controller tubes to the gas inlet of the breathing circuit |
| GUI | Computer | MacBook Pro, Apple | Any personal computer with Python installed can be used as computer. |
| | Software | Python program, Gustav Magnusson (author) | |
| | USB-RS485-converter | P/N USB-RS485-WE-5000-BT, FTDI Chip | USB converter which enables RS485 communication between computer and mass-flow controllers. |

**Table S3.** Products and manufacturers of components used in the Ventilator and Subject Setup breathing circuit.

| Component | | Product, Manufacturer | Comment |
|---|---|---|---|
| Ventilator Setup | Gas inlet | P/N 1947000 + 2710000, Intersurgical | A luer-connector for the mass-flow controller in Additional $CO_2$ System to connect to. |
| | Humidifier | MR290, Fisher & Paykel | An empty humidifier with no water, to mix gas. |
| | Connector humidifier | P/N 1992000, Intersurgical | Connecting gas inlet connector to humidifier. |
| | Coaxial ventilator tube | P/N Q712218N-IGS, Envisen | Standard ventilator tube, $2\,\mathrm{m}$ long. |
| | Elbow with sampling port | P/N 2714000, Intersurgical | To sample $O_2$/$CO_2$ gas. |
| | Test-lung | AccuLung, Fluke Biomedical | Test-lung with variable compliance $(10, 20$ and $50\mathrm{mL\,mbar^{-1}})$ and resistance $(5, 20$ and $50\mathrm{mbar\,L^{-1}\,s})$ |
| Subject setup | Expandable tube | P/N 1526000, Intersurgical | Expanded in Reservoir $CO_2$ setup, contracted in Additional $CO_2$ System. |
| | Gas inlet | P/N 1947000 + 2710000, Intersurgical | A luer-connector for the mass-flow controllers in Additional $CO_2$ or Reservoir $CO_2$ Systems to connect to. |
| | One-way valve | P/N 1921000, Intersurgical | Two used to control air flow. |
| | Filter | P/N M1003346, Timik Medical | Particle filter. |
| | Y-piece | P/N 1901000, Intersurgical | Separates inspiration/expiration side of circuit. |
| | Outlet adapter | P/N 1967000, Intersurgical | $22\,\mathrm{mm}$-female to $22\,\mathrm{mm}$ female adapter, to connect outlet of Y-piece to the one-way valve. |
| | Elbow with sampling port | P/N 2714000, Intersurgical | To sample $O_2$/$CO_2$ gas. |
| | Face mask adapter | 3D-printed, Gustav Magnusson (author) | $22\,\mathrm{mm}$-female to $30\,\mathrm{mm}$ male adapter. |
| | Face mask | Mask 7450 V2, Vyaire | Variable sized (XS to L). |

**Table S4.** The 14 ventilator test cases used to evaluate the Additional $CO_2$ method in mechanical ventilation. Note that the *Test case - Number* refers to the tables 201.104/201.105 in the European standard ISO 80601-2-12:2020.

| Test case | | Test lung parameters | | Ventilator settings | | | | | |
|---|---|---|---|---|---|---|---|---|---|
| Number | Control mode | Compliance ($\mathrm{mL\,mbar^{-1}}$) | Resistance ($\mathrm{mbar\,L^{-1}\,s}$) | Frequency (breaths/$\mathrm{min}$) | Insp. time ($\mathrm{s}$) | $FiO_2$ ($\%$) | PEEP (cmH2O) | Insp. pressure ($\mathrm{cmH_2O}$) | Tidal volume (L) |
| 1 | Volume | 50 | 5 | 20 | 1 | 30 | 5 | – | 500 |
| 2 | Volume | 50 | 20 | 12 | 1 | 90 | 10 | – | 500 |
| 3 | Volume | 20 | 5 | 20 | 1 | 90 | 5 | – | 500 |
| 4 | Volume | 20 | 20 | 20 | 1 | 30 | 10 | – | 500 |
| 5 | Volume | 20 | 20 | 20 | 1 | 30 | 5 | – | 300 |
| 6 | Volume | 20 | 50 | 12 | 1 | 90 | 10 | – | 300 |
| 7 | Volume | 10 | 50 | 20 | 1 | 30 | 10 | – | 300 |
| 1 | Pressure | 50 | 5 | 20 | 1 | 30 | 5 | 10 | – |
| 2 | Pressure | 50 | 20 | 12 | 1 | 90 | 10 | 15 | – |
| 3 | Pressure | 20 | 5 | 20 | 1 | 90 | 5 | 25 | – |
| 4 | Pressure | 20 | 20 | 20 | 1 | 30 | 10 | 25 | – |
| 5 | Pressure | 20 | 20 | 20 | 1 | 30 | 5 | 15 | – |
| 6 | Pressure | 20 | 50 | 12 | 1 | 90 | 10 | 10 | – |
| 7 | Pressure | 10 | 50 | 20 | 1 | 90 | 5 | 15 | – |
